# Multimodal Biomarker-Guided Deep Brain Stimulation Programming in Parkinson’s Disease: The DBSgram Framework

**DOI:** 10.64898/2026.03.29.26349663

**Authors:** Pedro Melo, Eduardo Carvalho, Andreia Oliveira, Ricardo Peres, Carolina Soares, Maria Rosas, Adriana Arrais, Rita Vieira, Duarte Dias, João PS Cunha, Manuel J. Ferreira-Pinto, Paulo Aguiar

## Abstract

Deep Brain Stimulation (DBS) is an effective therapy for Parkinson’s disease (PD), but clinical programming of stimulation parameters remains a time-consuming process largely guided by subjective symptom assessment. The increasing availability of sensing-enabled neurostimulators and wearable motion sensors provides an opportunity to introduce objective biomarkers into DBS titration. In this work, we present DBSgram, a multimodal framework designed to support data-driven DBS programming by integrating neurophysiological and kinematic measurements acquired during routine clinical titration. The proposed system combines subthalamic nucleus local field potential (STN-LFP) recordings from sensing-enabled neurostimulators with hand kinematic data acquired using wearable inertial measurement units (IMUs). A two-stage synchronization strategy aligns independent data streams from implanted and wearable devices, followed by automated signal processing pipelines for extracting electrophysiological and motor biomarkers. Patient-specific beta-band power is derived from LFP recordings, while tremor, rigidity, and bradykinesia metrics are computed from multi-axis IMU signals using symptom-specific processing algorithms. These synchronized features are then integrated into the DBSgram visualization framework, which maps stimulation amplitude to simultaneous changes in neural activity and objective motor performance. The framework was implemented in a standardized 40-minute clinical titration protocol conducted in a cohort of 18 PD patients implanted with sensing-enabled DBS systems.

We present here the analysis of aligned multimodal datasets from different patients to demonstrate proof-of-concept feasibility. The resulting DBSgram visualizations capture stimulation-dependent suppression of pathological beta activity alongside quantitative motor improvements, enabling intuitive identification of patient-specific therapeutic windows. These results demonstrate the technical feasibility of integrating implanted neurophysiological recordings with wearable kinematic sensing during DBS programming. By providing synchronized physiological and motor biomarkers within a unified framework, the DBSgram approach may support more objective and data-driven DBS titration, and contribute to future closed-loop neuromodulation strategies.

## Introduction

Deep Brain Stimulation (DBS) targeting the subthalamic nucleus (STN) is an established and effective therapy for the treatment of motor symptoms in Parkinson’s disease (PD), including tremor, rigidity, and bradykinesia [1], [2], [3], [4]. Despite its clinical success, the therapeutic outcome of DBS strongly depends on the appropriate selection of stimulation parameters, including amplitude, pulse width, frequency, and active contact configuration. In current clinical practice, this programming process remains largely empirical and relies on iterative adjustments guided by semi-quantitative clinical assessments such as the Unified Parkinson’s Disease Rating Scale (UPDRS) [5]. These evaluations require repeated patient examinations and are inherently subjective, making them susceptible to inter-rater variability and patient fatigue during prolonged programming sessions [6], [7]. As a result, DBS parameter optimization is often time-consuming and may lead to suboptimal stimulation settings or delayed identification of stimulation-induced adverse effects [5], [8].

Recent advances in neuromodulation technology have introduced new opportunities to support more objective DBS programming strategies. Sensing-enabled neurostimulators now allow the recording of local field potentials (LFPs) directly from implanted electrodes, enabling the monitoring of neural activity within the stimulated basal ganglia circuits [9]. In particular, excessive beta-band activity (13–35 Hz) in the STN has been consistently associated with motor impairment in PD and its suppression has been linked to clinical improvement during DBS therapy [4], [10], [11]. In parallel, wearable sensing technologies such as inertial measurement units (IMUs) have demonstrated the ability to capture quantitative measures of motor symptoms in PD patients [12], [13]. These wearable systems enable objective characterization of tremor, rigidity, and bradykinesia through continuous kinematic monitoring. Together, implantable neurophysiological recordings and wearable motion sensing provide complementary information that could support more data-driven approaches to DBS programming.

Despite these technological advances, integrating neurophysiological recordings and wearable motor measurements into practical DBS programming workflows remains a significant system-level challenge. In clinical settings, LFP recordings and wearable sensor outputs are typically acquired using independent hardware platforms with different acquisition protocols and internal clocks, which complicates their direct comparison and interpretation. Moreover, translating these heterogeneous data streams into clinically interpretable information requires robust synchronization strategies and automated signal processing methods capable of extracting reliable symptom-specific biomarkers. As a result, clinicians are often required to interpret multiple disjointed data sources rather than a unified representation of neural activity and motor performance. To address this limitation, a computational framework capable of synchronizing, processing, and jointly visualizing electrophysiological and kinematic signals (LFPs and IMUs) is required to support objective and quantitative DBS programming.

To address these challenges, we introduce **DBSgram**, a multimodal framework designed to integrate, synchronize, and visualize neurophysiological and motor data during DBS titration procedures. The DBSgram combines subthalamic nucleus LFP recordings obtained from sensing-enabled neurostimulators with kinematic measurements derived from wearable inertial sensors, enabling the simultaneous evaluation of neural biomarkers and objective motor metrics. By mapping stimulation amplitude to both beta-band activity and quantitative motor performance, the framework provides a unified representation of stimulation-induced physiological and behavioral changes. This integrated visualization aims to support DBS programming by facilitating the identification of patient-specific therapeutic windows and enabling a direct comparison between electrophysiological biomarkers and motor symptom improvements across stimulation conditions.

The main contributions of this work are threefold. First, we developed a multimodal acquisition framework that enables the synchronized recording of neurophysiological signals from sensing-enabled DBS implants and kinematic measurements from wearable inertial sensors during routine clinical titration procedures. A two-stage synchronization protocol was implemented to align independent data streams from implanted devices and wearable systems with sub-second accuracy. Second, automated signal processing pipelines were designed to extract objective symptom-related biomarkers from both modalities. Patient-specific beta-band power was derived from STN local field potential recordings, while tremor, rigidity, and bradykinesia metrics were computed from multi-axis IMU signals using symptom-specific signal descriptors. Third, these synchronized electrophysiological and kinematic features were integrated into the DBSgram, a visualization framework that maps stimulation amplitude to simultaneous changes in neural activity and motor performance. The framework was implemented within a standardized in-clinic titration protocol executed during routine outpatient sessions in a cohort of 18 PD patients. Analysis of a subset of synchronized datasets illustrates the feasibility of using multimodal biomarkers to visualize patient-specific therapeutic windows and support data-driven DBS programming

## Methods

The implementation of the proposed DBSgram consists of five primary stages: hardware integration, clinical data acquisition, multimodal signal synchronization, automated signal processing, and motor metrics mapping. The full pipeline is depicted in the block diagram presented in Fig.1. Initially, a standardized clinical titration protocol was conducted to collect simultaneous LFP and kinematic data from PD patients using sensing-enabled neurostimulators and wearable sensors. To ensure robust analysis, a two-level time-alignment procedure was applied to synchronize both data streams and a video feed. Subsequently, automated signal processing pipelines were used to extract symptom-specific “biomarkers” for tremor, rigidity, and bradykinesia from the multi-axis IMU data, while corresponding beta-band power was extracted from the STN-LFP recordings. Finally, these processed streams were integrated and mapped to generate the DBSgram. Data was processed and analysed using MATLAB R2025b.

**Figure 1.**
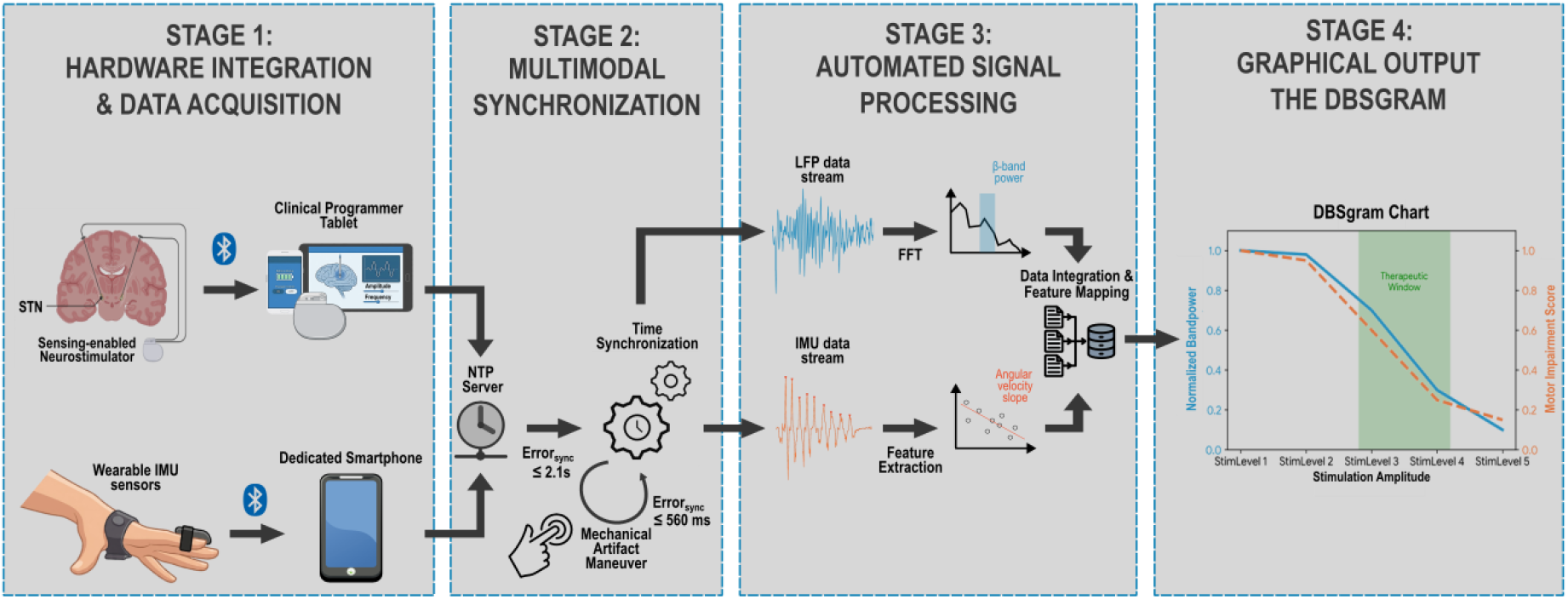
Block diagram illustrating the complete multimodal data acquisition and processing pipeline of the DBSgram framework. Stage 1 depicts the parallel recording of neurophysiological signals (STN-LFP) via the sensing-enabled neurostimulator and kinematic data via the wearable IMU sensors. Stage 2 details the robust two-level temporal synchronization protocol, utilizing a Network Time Protocol (NTP) server, in a first instance (Error_sync_ ≤ 2.1 s) and a mechanical artifact maneuver to align streams with sub-second accuracy (Error_sync_ ≤ 560 ms), on the offline analysis. Stage 3 outlines the automated, parallel signal processing pipelines for extracting the LFP beta-band power and objective motor metrics (e.g., angular velocity). Finally, Stage 4 demonstrates the integration of these synchronized features into one of the charts in the final DBSgram visualization to map the patient-specific therapeutic window.

### A. Participants and Hardware integration

An initial cohort of 18 patients with idiopathic PD undergoing STN-DBS was recruited. Inclusion criteria required a DBS therapy duration of less than 3 years and implantation with Medtronic’s Percept^™^ PC/RC neurostimulators featuring quadripolar directional leads (1-3-3-1 configuration), which allow for directional stimulation field steering. Participants were required to exhibit peak spectral power ≥1.2 μV^2^/Hz in the alpha-beta frequency band (8–35 Hz) on either lead while OFF stimulation or OFF medication (as per the guidelines of the ADAPT-PD clinical trial [14]). This study was approved by the Ethics Committee of Centro Hospitalar Universitário de São João (reference CE 233/2023), and all participants provided informed consent. Data were anonymized prior to analysis.

To capture objective motor metrics, bilateral hand kinematics were recorded by the iHandU wearable system [12], [15]. The setup used two “SnapKi” sensor modules per hand, each attached, respectively, to the index finger and palm (totalling four wearable sensors per session). Each module contains a Bluetooth-enabled 9-axis IMU, comprising a 3-axis accelerometer, gyroscope, and magnetometer. Each sensor pair transmits data wirelessly through Bluetooth to a smartphone for processing and visualization [16].

**Table 1.** Clinical Titration Protocol Sections.

| Section Number | Recording Duration (min) | Recording Type (<u>B</u>ilateral or <u>U</u>nilateral) | Description |
| --- | --- | --- | --- |
| 0 | 1 | B | Mechanical artifact generation (patient instructed to hit the neurostimulator's battery) for subsequent temporal alignment of data |
| 1 | 1 | B | Baseline neural activity recording with stimulation OFF |
| 2 | 10 (2×5) | U | Resting tremor study per hemibody with left (2L) and right (2R) hemisphere stimulation titration |
| 3 | 10 (2×5) | U | Wrist rigidity study per hemibody with left (3L) and right (3R) hemisphere stimulation titration |
| 4 | 10 (2×5) | U | Hand open-close study per hemibody with left (4L) and right (4R) hemisphere stimulation titration |
| 5 | 1 | B | Baseline neural activity recording with stimulation ON according to clinical prescription |

In parallel with the kinematic stream stored locally in the smartphones, the Percept^™^ system’s Brainsense^™^ modalities were used to acquire LFP data. The *Survey* and *Setup* modes were executed initially to identify prominent frequency peaks and validate signal integrity, while the *Streaming* mode acquired real-time, bilateral STN LFPs at 250 Hz during the clinical protocol [17], [18]. A clinician’s programmer tablet served as the primary interface with the patient’s implantable pulse generator (IPG) for parameter adjustment and data extraction. Upon session completion, all IPG data streams were exported directly from the tablet as a consolidated JSON file for offline analysis and second-stage temporal synchronization with the IMU data.

### B. Clinical Titration Protocol

An in-clinic titration protocol was carefully designed to balance clinical feasibility with robust data acquisition. While the dynamic nature of LFPs and fluctuating PD symptoms inherently introduce variability, the full protocol’s duration (∼40 minutes) was optimized to minimize the procedural burden on both patients and clinicians while ensuring sufficient continuous data for robust metric extraction.

The complete protocol comprised nine distinct recordings divided into six sections (labelled 0 to 5, summarized in Table I). Patients performed specific static (rest) and dynamic motor tasks (wrist rigidity and hand open-close) to evaluate the cardinal PD symptoms. Specifically, the goal was to also quantify the alignment with clinical scoring using the Movement Disorder Society (MDS)-Sponsored Revision of the UPDRS Scale Part III.

The protocol started with a 1-minute bilateral recording (Section 0) during which the patient generated a mechanical artifact by tapping the skin over the IPG at given intervals with a closed fist of the left hand, establishing the common marker for subsequent offline temporal alignment on all three data streams. This initial synchronization step was immediately followed by a 1-minute baseline neural activity recording with stimulation completely turned OFF (Section 1). The core of the clinical assessment then progressed through three pairs of 5-minute sections designed to evaluate the direct effect of varying stimulation amplitudes on the specified motor symptoms: resting tremor (MDS-UPDRS item 3.17; Section 2), wrist rigidity (item 3.3; Section 3), and hand bradykinesia (item 3.5; Section 4). These paired sections (e.g., 2L and 2R) correspond to the independent titration of each individual hemisphere.

During these symptom-evaluation sections, the titration paradigm began with bilateral 0 mA stimulation. Amplitude was subsequently increased unilaterally in 1-minute intervals. A total of four evenly spaced amplitude increments were applied, yielding five distinct 1-minute stimulation levels per hemisphere. These increments were predetermined such that the second to last step matched the clinician’s predefined optimal therapeutic level, allowing the final step to briefly test the upper bound of the therapeutic window. Crucially, these stimulation increments were applied exclusively to the hemisphere contralateral to the evaluated hemibody. Upon completing the 5-minute titration sequence for one hemisphere, the process was systematically repeated for the contralateral hemisphere, yielding, for example, separate 2L and 2R recordings for tremor.

After all motor tasks and amplitude titrations were completed, the session concluded with a final 1-minute bilateral baseline recording with stimulation turned ON at the clinician’s predefined optimal settings (Section 5).

### C. Multimodal Synchronization Pipeline

The temporal alignment of the three data streams is a critical system-level challenge when integrating independent clinical hardware. Because the Medtronic Percept^™^ neurostimulator, the iHandU wearable system and the video camera operate on independent internal clocks, we implemented a robust two-level synchronization methodology to establish a unified timeline for the DBSgram. The first level of synchronization relied on a Network Time Protocol (NTP) server. By binding the neurostimulator’s clinical programmer tablet and the iHandU receivers and video smartphones to the same NTP server via a local hotspot, we achieved a preliminary, coarse synchronization with an error margin of < 2.1 seconds across all data streams [19]. To achieve the fine-grained alignment necessary for precise physiological mapping, a secondary mechanical artifact maneuver was employed. As described in Section B, patients were instructed to physically tap the implanted pulse generator using their sensor-equipped hands at the beginning of the recording sessions while in view of a clinical camera. This maneuver generated a distinct, simultaneous mechanical marker in the IMU accelerometer data, the LFP recordings (as a motion artifact), and the video feed. During offline downstream analysis, these multi-domain markers were identified and temporally aligned, successfully reducing the final synchronization error to ≤ 560 ms [19]. This rigorous alignment ensured that transient neurophysiological fluctuations could be reliably correlated with instantaneous kinematic responses.

### D. Kinematic Signal Processing

To translate raw multi-axis IMU data into objective clinical metrics, data continuously acquired from the 16-bit wearable sensors at a sampling rate of 50 Hz were fed into automated signal processing pipelines implemented for each evaluated cardinal symptom.

For tremor analysis, accelerometric signals acquired from the finger sensors were first filtered using a 6th-order band-pass Butterworth filter with cutoff frequencies of 2 and 8 Hz, to isolate the characteristic frequency band for Parkinsonian tremor [20], [21]. The filtered total acceleration was segmented into 3-second epochs with a 50% overlap. To distinguish genuine tremor from rest or voluntary movement, a decision tree algorithm was applied based on spectral power distribution. For each epoch, spectral power was calculated across a tremor band (4–7 Hz), a low-frequency band (2–3 Hz), and a total band (2–8 Hz). Epochs exhibiting a root mean squared (RMS) acceleration of less than 0.003 m/s^2^ were strictly labeled as rest. Epochs were classified as tremor if the ratio of tremor-band power to low-frequency power exceeded 6.5, or if the ratio exceeded 2.0 while tremor dominance (tremor-band power divided by total-band power) was simultaneously ≥50%. Temporal continuity rules were subsequently applied to merge adjacent tremor segments and relabel isolated, fragmented epochs. For all confirmed tremor epochs, the maximum RMS acceleration value was extracted to serve as a continuous, objective correlate to the clinical UPDRS tremor score. While highly robust for pronounced symptoms, it is noted that this accelerometric thresholding exhibits limitations in reliably distinguishing extremely mild tremors (lower-bound UPDRS score of 1) from the baseline resting state, as the mechanical deviations approach the sensor’s noise floor.

Wrist rigidity was assessed by evaluating the angular velocity recorded by the IMU’s gyroscope during the standardized passive flexion-extension maneuvers. The motor data were fed into a previously validated, pre-trained computational model. This model, developed using a rigorously labeled 45-patient dataset (comprising 403 discrete gyroscopic signals), evaluates patented signal descriptors to output a rigidity improvement percentage relative to a UPDRS-4 baseline severity [15]. When evaluated against an independent 14-patient clinical test set (344 gyroscopic signals), this approach allowed for the direct quantification of rigidity with an established accuracy of 82.0% and an average test error of 3.4%, eliminating reliance solely on the physician’s subjective tactile perception [15].

Finally, hand bradykinesia was quantified using data from the repetitive hand open-close tasks. The raw gyroscopic signals were band-pass filtered between 1 and 20 Hz to eliminate high-frequency noise and baseline drift. The signals were then segmented to identify individual open and close cycle peaks. To capture the progressive decrement in speed and amplitude characteristic of Parkinsonian fatigue, linear interpolation was applied to the peaks of the filtered angular velocity, specifically extracting the main component of the rotation axis from the finger sensor across the exercise duration. From these segmented cycles, two primary metrics were extracted: the total signal power, representing the overall kinetic energy expended during the task, and the slope of the angular velocity. The slope served as the primary descriptor of motor fatigue; a steeper negative slope indicated a more rapid decline in motor performance [10].

### E. Neurophysiological Signal Processing

To complement the objective kinematic metrics, simultaneous LFP recordings acquired directly from the STN were processed to extract corresponding neurophysiological biomarkers. Initial calibration data, obtained via the neurostimulator’s *Survey* and *Setup* modalities prior to the protocol, were segmented into 250-sample (1-second) windows. To ensure high spectral resolution and mitigate signal artifacts, these segments were systematically detrended, windowed using a Hann function to reduce spectral leakage, and zero-padded before executing a 512-point Fast Fourier Transform (FFT). The resulting normalized power spectra were used to identify patient-specific peak frequencies within the beta band, establishing the baseline neurophysiological profile.

During the continuous clinical titration protocol, the real-time *Streaming* data were segmented into 3-second epochs explicitly designed to match the temporal boundaries of the movement states (e.g., rest, tremor, or voluntary movement) established by the automated IMU processing pipeline. For the rigidity and bradykinesia assessments, periods of active movement were identified through acceleration thresholding and further corroborated using the synchronized clinical video recordings. To accurately quantify the physiological impact of the neuromodulation therapy, the absolute beta-band power for both tremor and resting states was calculated for each discrete epoch. These raw power values were then normalized to the median beta power of all resting epochs recorded at the lowest tested stimulation level (typically 0 mA). This standardized normalization procedure facilitated the robust, relative quantification of stimulation-driven beta suppression across the entire amplitude titration spectrum.

### F. Data Quality and Exclusion Criteria

To ensure the proof-of-concept validation of the DBSgram framework, all acquired recordings were subjected to rigorous data quality and exclusion filters. The primary objective was to eliminate confounding variables that could compromise the accuracy of the multimodal temporal alignment or the integrity of the extracted clinical biomarkers. Consequently, datasets were excluded if a patient exhibited clinical intolerance that prevented the completion of all predefined titration amplitude steps, or if the contralateral stimulation was inadvertently left active while isolating a specific hemisphere’s therapeutic response.

Furthermore, recordings were discarded if persistent, broad-band stimulation interference or hardware motion artifacts significantly contaminated the LFP frequency spectra during a continuous titration sequence. On the wearable sensors side, sessions characterized by excessive, non-tremor voluntary movements were removed, as these continuous interruptions precluded the computation of a reliable, stable resting baseline necessary for the beta-band power normalization. Following the application of these strict data quality filters and the mandatory verification of sub-second multimodal synchronization (as detailed in Section II.C), complete and precisely aligned datasets were successfully retained for a subset of 4 patients. This rigorous curation yielded 8 fully mapped and highly reliable hemispheres for the final DBSgram system demonstration and analysis.

## Results

### A. Data Acquisition and Dataset Characteristics

The proposed multimodal framework was deployed during routine clinical DBS titration sessions conducted in a cohort of 18 patients with Parkinson’s disease implanted with sensing-enabled neurostimulators. The standardized protocol required approximately 40 minutes per patient and was successfully completed without disrupting the normal outpatient workflow.

Following data acquisition, all recordings were evaluated using the predefined synchronization and signal quality criteria described in Section II. Datasets were excluded if multimodal alignment could not be reliably established, if stimulation artifacts severely contaminated the LFP recordings, or if excessive voluntary movements prevented the extraction of stable motor baselines.

### B. Multimodal Signal Synchronization

The proposed two-stage synchronization pipeline successfully aligned LFP recordings, wearable sensor data, and video recordings acquired from independent hardware platforms. Initial coarse synchronization using a Network Time Protocol (NTP) server provided temporal alignment between acquisition devices with an error margin below 2.1 seconds.

Subsequent fine synchronization was achieved using the mechanical artifact maneuver described in Section II.B, which generated a distinct marker simultaneously observable in the IMU accelerometer signals, LFP recordings, and video stream. Offline alignment of these markers reduced the synchronization error to ≤ 560 ms across data streams. This level of temporal precision enabled the association of electrophysiological fluctuations with the corresponding motor states identified from the wearable sensors during the titration protocol.

### C. Biomarker Extraction from LFP and IMU Signals

The automated signal processing pipelines successfully translated raw IMU and LFP data into continuous, objective metrics, respectively, of the patient’s motor response and electrophysiological state. Across the analysed proof-of-concept datasets, the system demonstrated a robust capacity to simultaneously capture physiological suppression and symptomatic motor changes.

From a neurophysiological perspective, the framework accurately tracked stimulation-induced alterations in pathological STN oscillations. As observed in the representative “ideal responder” profile, detailed in Fig. 2, the LFP processing pipeline reliably isolated patient-specific beta-band activity. During the unilateral stimulation titration sequence, the relative beta-band power exhibited a consistent, dose-dependent decrease as the stimulation amplitude was incrementally increased from the 0 mA baseline (Stim OFF) up to 2.0 mA. This specific neurophysiological response confirms the system’s ability to accurately quantify the progressive dampening of the hypersynchronous STN network activity in real-time.

**Fig. 2.**
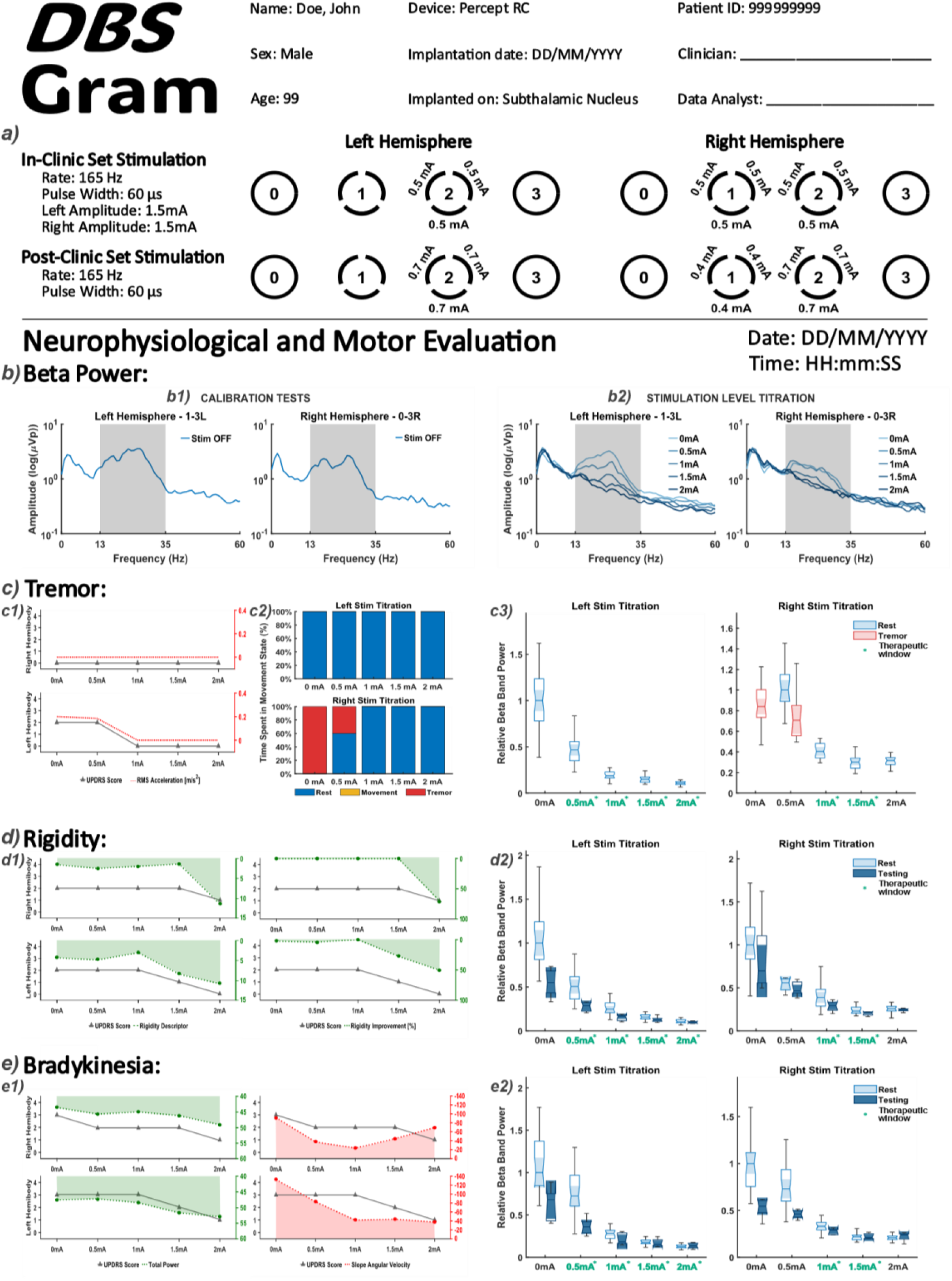
Representative DBSgram clinical report for Patient 007. The multimodal visualization integrates (**a**) clinical stimulation parameters, (**b**) LFP beta-band power spectra, and (**c–e**) parallel evaluations of cardinal motor symptoms, mapping clinical UPDRS subscores (grey) against objective kinematic metrics (colored). For the left hemisphere, the framework captures a highly correlated, dose-dependent suppression of both pathological beta-band power and motor impairment, clearly delineating a broad and stable therapeutic window (0.5 to 2.0 mA, teal green numbers with asterisk). Based on this quantitative confirmation, the final titration level on the left hemisphere was selected as the new therapeutic amplitude without adverse effects. Furthermore, the right hemisphere titration highlights the tool’s utility in advanced programming: while the highest amplitude (2.0 mA) yielded optimal physiological and kinematic improvements, it simultaneously induced capsular side effects. Guided by the DBSgram’s objective confirmation of maximum therapeutic efficacy at this threshold, the clinician successfully utilized directional steering to redistribute the electrical field, preserving the optimal motor response while safely mitigating the adverse effects.

Concurrently, the kinematic processing pipelines successfully quantified the corresponding motor symptom variations. The automated algorithms translated raw, multi-axis IMU data into symptom-specific descriptors. When evaluating these metrics for Patient 007, the continuous, sensor-derived descriptors, in particular, for this case, the RMS acceleration for resting tremor (Fig. 2c1) and the computational angular velocity evaluations for wrist rigidity (Fig. 2d1) and hand bradykinesia (Fig. 2e1), closely mirrored the stepwise improvements documented by the clinician-rated MDS-UPDRS Part III sub-scores. Notably, the parallel decline also observable in the pathological beta-band power (Fig. 2c3, Fig. 2d2, Fig. 2e2) robustly validates the framework’s capability to directly link underlying pathophysiology with physical motor execution.

The successful, synchronized extraction of these multi-domain biomarkers formed the foundational data layer required for the generation of the comprehensive DBSgram visualizations.

### D. DBSgram Visualization and Therapeutic Window Identification

By synchronizing and co-registering these independent data streams, the DBSgram framework successfully generated comprehensive clinical reports, facilitating the personalization of DBS therapies. To illustrate the clinical utility of this proposed visualization tool, varying titration profiles from two distinct patients in the validated cohort are presented (Figs. 2 and 3), ranging from “ideal responder” hemispheres to more complex ones.

**Fig. 3.**
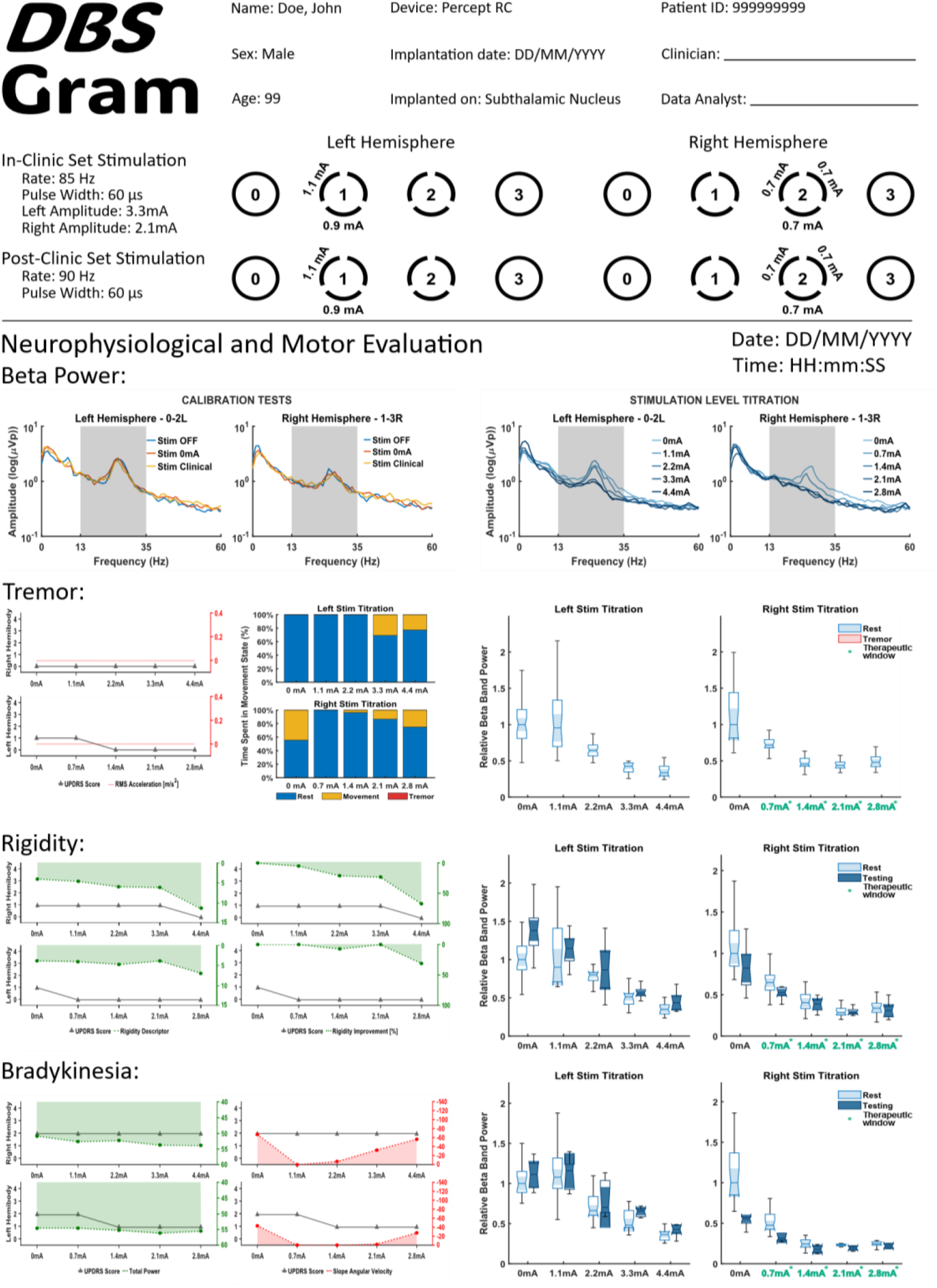
Representative DBSgram clinical report for Patient 014 demonstrating a severe intra-patient asymmetry of neurostimulator programming. The multimodal visualization similarly integrates clinical stimulation parameters, LFP beta-band power spectra, and parallel evaluations of cardinal motor symptoms, mapping UPDRS subscores (grey) against objective kinematic metrics (colored). The right hemisphere (evaluating the left hemibody) exhibits an ideal, highly responsive profile, capturing a continuous, highly correlated suppression of beta power and motor impairment to yield a broad, side-effect-free therapeutic window (0.7 to 2.8 mA). In stark contrast, the left hemisphere reveals a significantly delayed therapeutic response. UPDRS scores and kinematic metrics for rigidity and bradykinesia remain stagnant across the lower amplitudes (0 to 3.3 mA), despite a concurrent, progressive decrease in beta-band power. Clear motor improvement is only exhibited at the highest tested level (4.4 mA). However, the simultaneous onset of stimulation-induced adverse side effects at 4.4 mA results in an absent therapeutic window across these coarse testing intervals. By visually mapping this delayed improvement against the beta-power curve, the DBSgram alerts the clinician that high-resolution amplitude testing is strictly required specifically between the pre-defined clinical setting (3.3 mA) and the upper boundary (4.4 mA) to safely optimize therapy.

The framework’s baseline utility is demonstrated in the former, prototypical, highly responsive hemibodies, specifically the left hemisphere of Patient 007 (Fig. 2) and the right hemisphere of Patient 014 (Fig. 3). In both instances, the DBSgram captures a smooth, highly correlated, and dose-dependent suppression of pathological beta-band power that directly aligns with steady improvements in objective kinematic symptom metrics. For Patient 014’s right hemisphere, UPDRS scores for rigidity and bradykinesia drop to zero by 1.4 mA, establishing a broad, stable therapeutic window ranging from 0.7 to 2.8 mA. Similarly, Patient 007’s left hemisphere exhibits a stable window from 0.5 to 2.0 mA. In these straightforward scenarios, the framework acts as a rigorous, quantitative confirmation of the clinician’s standard visual assessment, empowering physicians to confidently and rapidly establish optimal therapeutic amplitudes.

Interestingly, true clinical value of the DBSgram, however, is revealed when navigating complex, nuanced programming challenges, as seen in the remaining hemispheres of these same patients. The evaluation of the right hemisphere in Patient 007 presents a scenario where the highest tested amplitude (2.0 mA) yielded the most robust physiological and kinematic improvements but simultaneously triggered stimulation-related capsular adverse side effects. Because the DBSgram provided objective confirmation that 2.0 mA offered the absolute maximum therapeutic efficacy, the clinician was justified in targeting this higher amplitude. Guided by the data, the clinician successfully utilized directional steering to redistribute the electrical field, preserving the optimal motor response while safely mitigating the adverse effects (see Fig. 2a).

Conversely, the left hemisphere of Patient 014 (Fig. 3) reveals a significantly delayed therapeutic response. Both the UPDRS scores and objective kinematic metrics remain completely stagnant across the lower amplitude increments (0 to 3.3 mA), despite a concurrent, progressive decrease in beta-band power. Clear motor improvement is only exhibited at the highest tested level (4.4 mA). However, the simultaneous onset of stimulation-induced adverse side effects at this exact increment creates a clinical paradox, resulting in an absent therapeutic window across these coarse testing intervals. By visually contrasting this delayed motor improvement against the continuous biological beta-power suppression, the DBSgram alerts the clinician that the underlying neural circuits are responding, and that high-resolution amplitude testing (e.g., fractional milliampere steps) is strictly required between 3.3 mA and 4.4 mA to safely optimize therapy. Note here that the clinician until further testing, opted to end the session on its former predetermined therapeutical level, but opted to refine it after the protocol was implemented.

Overall, these results demonstrate the feasibility of integrating multimodal neural and kinematic biomarkers within a unified visualization framework to support objective DBS programming.

## IV. Discussion

The optimization of DBS therapy remains a significant clinical bottleneck, heavily reliant on time-consuming trial-and-error programming and subjective visual assessments. In this study, we introduced the DBSgram, designed to directly address this gap by seamlessly integrating neurophysiological biomarkers with IMU sensor-derived kinematic metrics.

The results from our proof-of-concept subset demonstrate the technical feasibility and clinical utility of this multimodal approach. By successfully executing the ∼40-minute in-clinic titration protocol, we demonstrated that robust, synchronized data can be acquired without disrupting the clinical outpatient workflow. Furthermore, the contrast between and within the analyzed patient profiles highlights the core value of the tool. In prototypical cases, such as the “ideal responder”, the framework serves as a rigorous, quantitative confirmation of the clinician’s visual assessment, confirming that physiological beta-suppression directly correlates with objective motor improvements. Crucially, the DBSgram also provides the objective confidence required for advanced programming interventions; as demonstrated, it can justify the use of directional steering to mitigate capsular side effects while preserving optimal motor relief.

However, the framework’s true clinical utility is further revealed in complex cases, such as the one in Fig. 3. By visually mapping delayed motor responses against the physiological curve and having adverse side-effect thresholds defining therapeutic window limits, the DBSgram effectively transforms disjointed data streams into an actionable clinical dashboard. Additionally, by instantly visualizing severe intra-patient asymmetry, the tool empowers physicians to allocate their time efficiently, applying rapid programming to a highly responsive hemisphere while dedicating high-resolution titration to a more resistant, narrow-window contralateral side.

Despite these promising preliminary outcomes, several limitations must be acknowledged. First, to ensure the integrity of the multimodal synchronization and the accuracy of the automated biomarker extraction, rigorous data quality filters were applied. This strict curation resulted in a heavily filtered subset of 4 patients from the initial cohort of 18. Datasets were frequently excluded due to persistent stimulation-induced hardware artifacts in the LFP recordings or non-task-related voluntary movements that prevented the establishment of a stable resting baseline. Consequently, while the current algorithms perform robustly on clean data, the signal processing pipelines require further optimization to handle the real-world environments inherent to routine clinical practice. Furthermore, this study was conducted exclusively in a clinical setting, which may not fully capture the complex, fluctuating nature of Parkinsonian symptoms during a patient’s daily life.

Future work will focus on expanding the validation cohort to statistically quantify the correlation between the DBSgram metrics and standard MDS-UPDRS outcomes across a broader Parkinsonian population. Additionally, algorithmic refinements will be prioritized to improve artifact rejection and dynamic state classification. Ultimately, translating this framework from an acute, in-clinic titration tool to a continuous monitoring system could provide clinicians with unprecedented insights into at-home symptom fluctuations, paving the way for highly personalized, data-driven, and potentially adaptive DBS (aDBS) programming paradigms.

## V. Conclusion

This study presents the DBSgram, a novel, multimodal tool whose goal is to objectify and streamline Deep Brain Stimulation (DBS) programming for Parkinson’s disease. The DBSgram aims to replicate in DBS the importance of an electrocardiogram or audiogram has in their respective fields. By successfully synchronizing neurophysiological STN-LFP recordings with continuous kinematic wearable sensors data, we demonstrated the technical feasibility of extracting and co-registering symptom-specific biomarkers during routine clinical titration. The preliminary proof-of-concept results validate that the automated metrics track closely with clinical MDS-UPDRS evaluations, effectively capturing both the physiological suppression of pathological beta-band activity and the concurrent improvement in motor execution. Crucially, the resulting visual dashboard translates complex, multi-domain data into an intuitive clinical report, allowing physicians to clearly delineate patient-specific therapeutic windows and identify nuanced thresholds for adverse side effects and confidently guide advanced interventions such as directional steering. While future large-scale validation and algorithmic refinement are necessary to handle real-world clinical noise and establish its broader clinical applicability, the DBSgram represents a significant step toward data-driven, personalized neuromodulation, laying the quantitative groundwork for more efficient and objective DBS therapy management.

## Data Availability

All data produced in the present study are available upon reasonable request to the authors

## Acknowledgments

This work was supported by Prémio Mantero Belard, Santa Casa da Misericordia de Lisboa (grant MB-12-2022), and by Foundation ‘la Caixa’ - CaixaResearch Health 2022 (grant HR22-00189).

